# Real-world experience with lecanemab therapy for Alzheimer’s disease in the Intermountain West

**DOI:** 10.1101/2025.09.30.25337002

**Authors:** Bethany C. Curd, Camilla Zubrick, Christine J. Cliatt Brown, Michelle Keown Sorweid, Sarah B. Dehoney, Yoshimi Anzai, Satoshi Minoshima, Anna L. Parks, Nicholas A. Frost

**Author notes:** Corresponding Author Correspondence: Dr. Nicholas A. Frost, Assistant Professor University of Utah Department of Neurology, Salt Lake City, UT @nickfrostneuro.

## Abstract

**INTRODUCTION:** Lecanemab is a monoclonal antibody targeting amyloid plaques that has been approved for the treatment of Alzheimer’s disease. Here, we report on the clinical history and outcomes of the first 70 patients at the University of Utah to receive amyloid-removal therapy.

**METHODS:** This is a retrospective analysis of patients treated with lecanemab over a 26-month period. We extracted patient data from charts and analyzed demographics, health history, and clinical details with outcomes on lecanemab treatment.

**RESULTS:** In total, we observed 14 cases of ARIA, which was significantly associated with ApoE genotype. Zero cases of ARIA were symptomatic, and there was no association between geographical location and adverse effects.

**DISCUSSION:** Our study examined the safety and tolerability of centralized lecanemab administration across a widely distributed region and suggests that use of distributed infusion sites increases access to disease-modifying treatment without significant increase in risk.

## BACKGROUND

Alzheimer’s disease (AD) is a highly prevalent^1-3^ and progressive neurodegenerative disorder^4,5^ marked by early deficits in spatial processing^6^ and memory. The pathological hallmark of the disease is amyloid plaques^7-10^ and tangles of aggregated tau^11^, which accumulate in the brains of individuals well before appreciable changes in cognition^12-15^. The stereotyped sequence of AD-associated changes led to the development of a model in which pathologic accumulation of amyloid creates a permissive environment for phosphorylation of tau and cellular death^16^. Thus, logically, removal of amyloid should slow down the progression of neurodegeneration in AD.

This was demonstrated in two large clinical trials involving monoclonal antibodies targeting aggregated amyloid-beta (Aβ) in the brains of patients with AD^7,17^. These trials led to the FDA approval of lecanemab in 2023 and donanemab in 2024 for treatment of cognitive decline in patients with mild cognitive impairment (MCI)^18^ or mild dementia due to AD.

Although these successful trials represent a tremendous advance for patients, there is little data available regarding their use in real-world settings. Outside the carefully controlled clinical trial setting, there are a number of logistical issues related to delivery that patients^19^ and hospital systems must navigate. Moreover, the long-term benefit must be balanced against the up-front risk of serious side effects, including infusion reactions and amyloid-related imaging abnormalities (ARIA), which occur in patients with AD but are more frequent in those receiving amyloid-removing medications^7,17,20-23^.

The University of Utah is the only university health system in the state of Utah and in the Intermountain West and provides care to patients within a 1.4 million-square-kilometer region encompassing Utah, Idaho, Montana, Nevada, and Wyoming. This large catchment area provides unique challenges related to providing care in geographically distributed and rural regions, particularly for patients with cognitive impairment.

In this study, we examined data collected from patients who were prescribed lecanemab at the University of Utah between April 2023 and July 2025. This initial cohort included patients from seven states. All patients were evaluated in cognitive neurology or geriatrics clinics and underwent neurological evaluation, magnetic resonance imaging (MRI), and biomarker confirmation via cerebrospinal fluid (CSF) or amyloid positron emission tomography (PET).

Patients were discussed at a multidisciplinary consensus conference and underwent surveillance MRIs and follow-up visits (in-person or via telehealth) to monitor for adverse events. We examined the rate of ARIA and other adverse events, discontinuation of therapy, time required to complete the first 13 and 26 doses of therapy, and by location relative to the clinical center of the University of Utah.

## METHODS

Under an Institutional Review Board-approved protocol and according to the Declaration of Helsinki, we performed a retrospective review of the first 70 patients treated with lecanemab at the University of Utah in Salt Lake City, Utah. Patients were evaluated in cognitive neurology or geriatrics clinics between April 2023 and July 2025. Three providers—two in neurology (N.A.F. & C.J.C) and one in geriatrics (M.K.S.)—provided cognitive evaluations consisting of an in-person neurological examination, MRI imaging, and biomarker confirmation with CSF evaluation or amyloid PET. Most patients also underwent advanced molecular imaging (i.e., metabolic PET) or molecular diagnostics (plasma p-tau217).

Inclusion criteria for initiation of lecanemab largely adhered to published appropriate use recommendations^24^ and included a diagnosis of MCI or mild dementia with biomarker confirmation of amyloid pathology, apolipoprotein E (ApoE) genotyping, and a baseline MRI within one year. While patients received counseling on risks of ARIA, including ApoE genotype, ApoE genotype was not an exclusionary criterion. Patients were deemed ineligible for therapy if actively using anticoagulants, exhibiting contraindications on MRI scans, or having an existing diagnosis of cerebral amyloid angiopathy (>4 microhemorrhages or a single microhemorrhage >1 cm in diameter), superficial siderosis, or other medical or psychiatric conditions thought to put them at greater risk of complications.

Patients who met inclusion criteria were counseled on the risks and benefits of therapy and prescribed infusions of lecanemab dosed at 10 mg/kg body weight. For safety and to ensure MRIs were completed and reviewed at recommended intervals, lecanemab was prescribed only for the number of doses specified between MRIs, with each sequence of infusions prescribed after MRI completion. Patients received surveillance MRIs after doses 4, 6, 13, 26, and 38 (and prior to receiving the subsequent dose) to monitor for subclinical changes on imaging (ARIA). Patients also received other therapeutics (e.g., Donepezil) as appropriate. Post-infusion MRI findings were graded for ARIA-E or ARIA-H per criteria in **Supplementary Table 1-A**. The response to MRI findings or symptoms depended on the severity, as detailed in **Supplementary Tables 1-B and 1-C.**

Demographic and clinical data was evaluated in a retrospective manner, extracted from patient charts using a standardized data abstraction form. We calculated distance from the clinic as the straight-line distance between the patient’s zip code and the Clinical Neurosciences Center at the University of Utah. Dividing patients into groups of those within 75 kilometers and those more than 75 kilometers from the clinic, we examined whether greater distance from the clinic might lead to differences in time to initiating or completing therapy or to disparities in clinical outcomes.

We next evaluated other factors that might influence or predict the rates of ARIA in our population. As part of cognitive care, patients underwent neurological exams, Montreal Cognitive Assessment (MoCA), Functional Activity Questionnaire (FAQ), ApoE genotyping, MRI imaging, and biomarker confirmation of amyloid pathology. We systematically collected individual patient medical history—including comorbidities, medication use, cognitive assessment scores, suspected co-pathologies, mean arterial blood pressure (MAP), body mass index (BMI), and ApoE genotype, as well as results of MRI, PET, lumbar puncture, and plasma testing—that we tallied and analyzed alongside ARIA case data to investigate any possible associations or confounders. Comorbidities included any specified by provider notes or medical history in patient charts and were totaled to assess relative prevalence. Any conditions occurring in three or fewer patients were removed from analysis. We reviewed charts for any active antiplatelet medications listed—including aspirin, clopidogrel, ticagrelor, cilostazol, and dipyridamole—again to examine the possibility of any associations with ARIA incidence.

Most patients also had metabolic PET, which allowed us to evaluate for potential co-pathologies. Limbic-predominant age-associated TDP-43 encephalopathy (LATE) is a highly prevalent driver of memory loss and neuropsychiatric changes in older adults^25-28^ and co-occurs in roughly 50% of patients with neuritic amyloid plaques^29^. We identified patients with possible LATE co-pathology based on metabolic PET images^27,30^ acquired during clinical evaluation. Specifically, we identified patients with significant hypometabolism affecting the medial temporal and inferior frontal lobes, as visualized by quantitative statistical maps^31^.

MoCA and FAQ were administered by trained medical assistants as part of clinical visits. We gathered each patient’s baseline scores before starting lecanemab treatment, as well as any subsequent scores completed since starting infusions. Similarly, we documented baseline blood pressure and most recent pre-infusion BMI. We stratified patient data from each category into subgroups for both the total patient group (n=70) and ARIA cases (n=14), analyzing each variable for potential relationships with ARIA outcomes. As ApoE genotype is known, based on anti-amyloid therapy clinical trials, to correlate with relative risk for ARIA, we grouped patient genotype results according to number of copies of the ApoE ε4 allele and measured the frequency against ARIA cases.

Baseline and surveillance MRI scans were reviewed for microhemorrhage, superficial siderosis, small vessel ischemic disease, and edema. Fazekas scores were calculated visually for each patient based on baseline MRI images as defined by Franz Fazekas, using the following criteria for deep white matter changes: Fazekas score 0 = absent; 1 = punctate foci; 2 = beginning confluence; 3 = large confluent areas.

Given the length of the study period and rolling nature of patient treatment timelines, we examined clinical history and outcomes against individual patient infusion history, characterizing each patient’s treatment course beginning with lecanemab referral date, first infusion date, total infusions as of July 1, 2025, and cases of discontinuing therapy.

We performed several statistical tests on the data, including Fisher’s exact test, Welch’s t-test, and two-way ANOVA with Bonferroni’s multiple comparisons. All analyses were performed in Prism 10 using two-tailed tests for all comparisons unless otherwise specified.

## RESULTS

During the period from July 2021 until April 2025, a total of 2372 patients were evaluated at the University of Utah by specialists in cognitive neurology (N.A.F. or C.J.C) or geriatrics (M.K.S.). The first eligible patients began receiving referrals for lecanemab infusions in April 2023. Of these, 70 patients had received at least one dose of lecanemab as of July 1, 2025, with 1222 doses of lecanemab administered in total during this period.

Over the course of treatment, 14 patients (20%) were observed to have radiographic features of ARIA-H, and two of these patients also had imaging showing ARIA-E (**Table 1**). The majority of these cases (71%) met criteria for mild ARIA, with three cases and one case classified as moderate and severe ARIA, respectively. Imaging findings were predominantly observed within the first six months of therapy (in 13 out of 14 cases), with only one patient demonstrating ARIA-H after dose 26. Two patients experienced two separate incidences of ARIA. No patients had symptomatic ARIA.

**Table 1:**
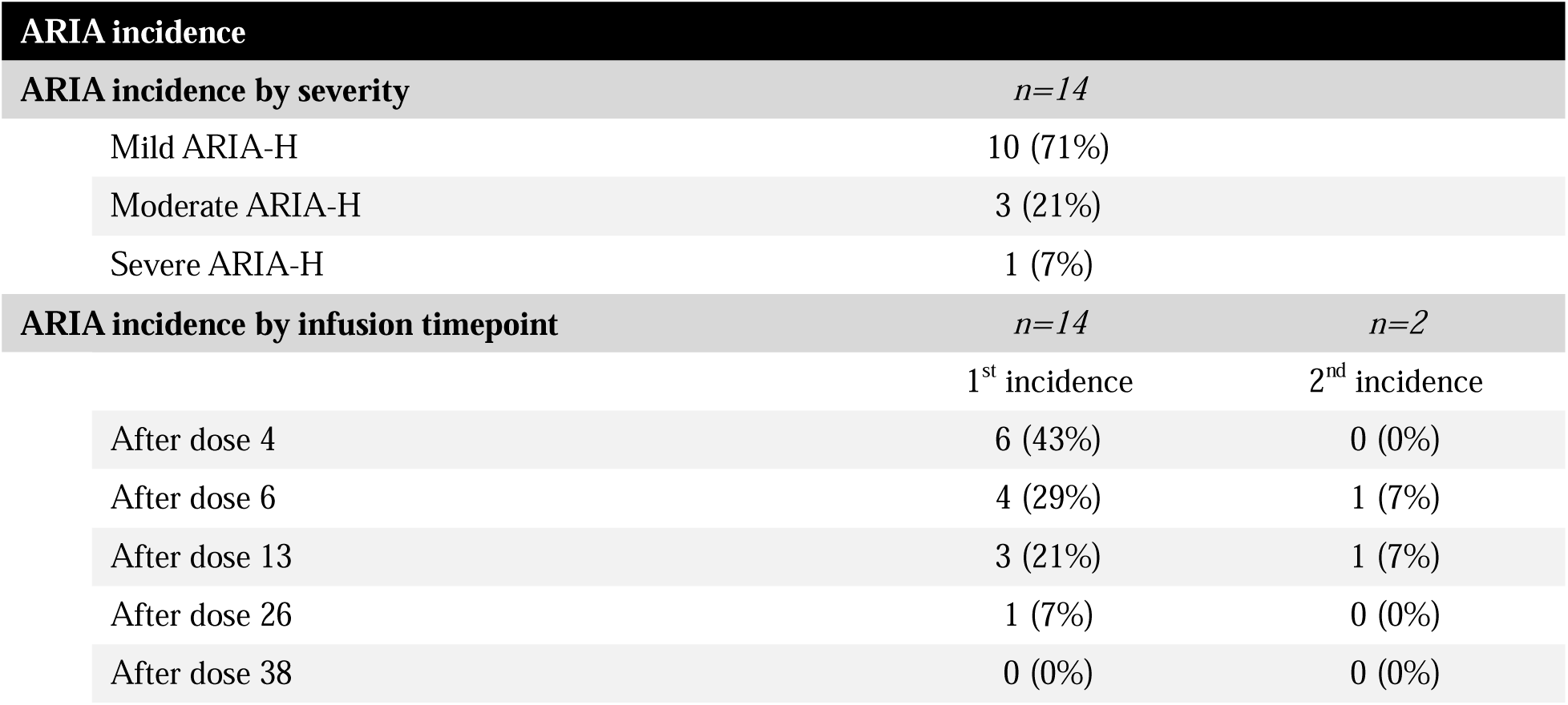
ARIA incidence. The relative severity of ARIA cases, as well as when the cases were identified during treatment, is included here. For ARIA incidence by severity, percentages represent the share within the ARIA cases category (n=14). For ARIA incidence by infusion timepoint, percentages in both columns also represent the relative fraction of total ARIA cases (n=14).

Of the total 70 patients in our study, 39 (56%) were male and 31 (44%) were female. The average age at first visit was 72.5, with no significant difference in mean age—either age at first visit or current age—between male and female patients. Two patients were Hispanic/Latino and one patient chose not to disclose; the remainder identified as White/non-Hispanic. We observed no difference in ARIA incidence according to sex, age, or race/ethnicity (**Table 2**).

**Table 2:**
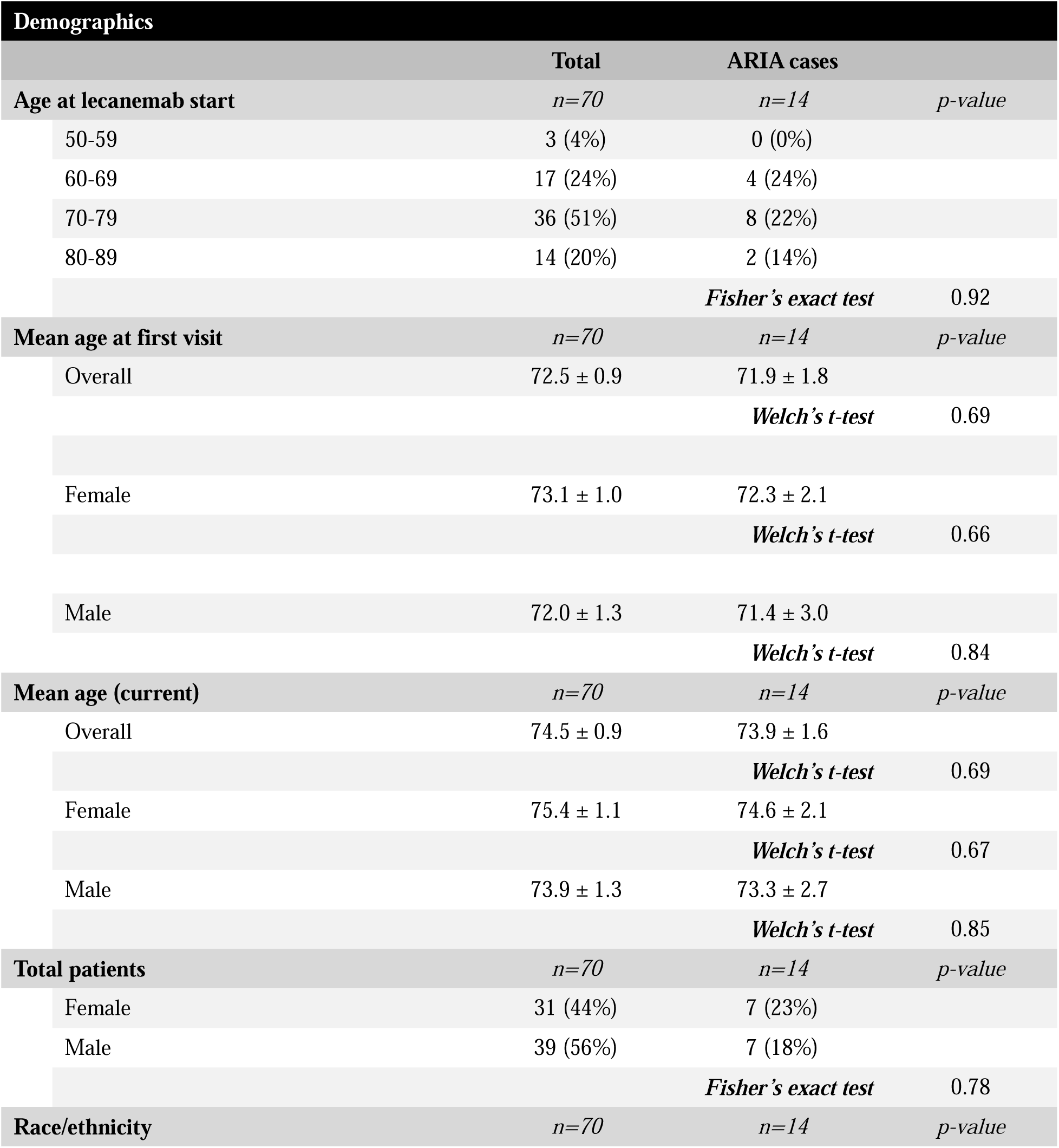
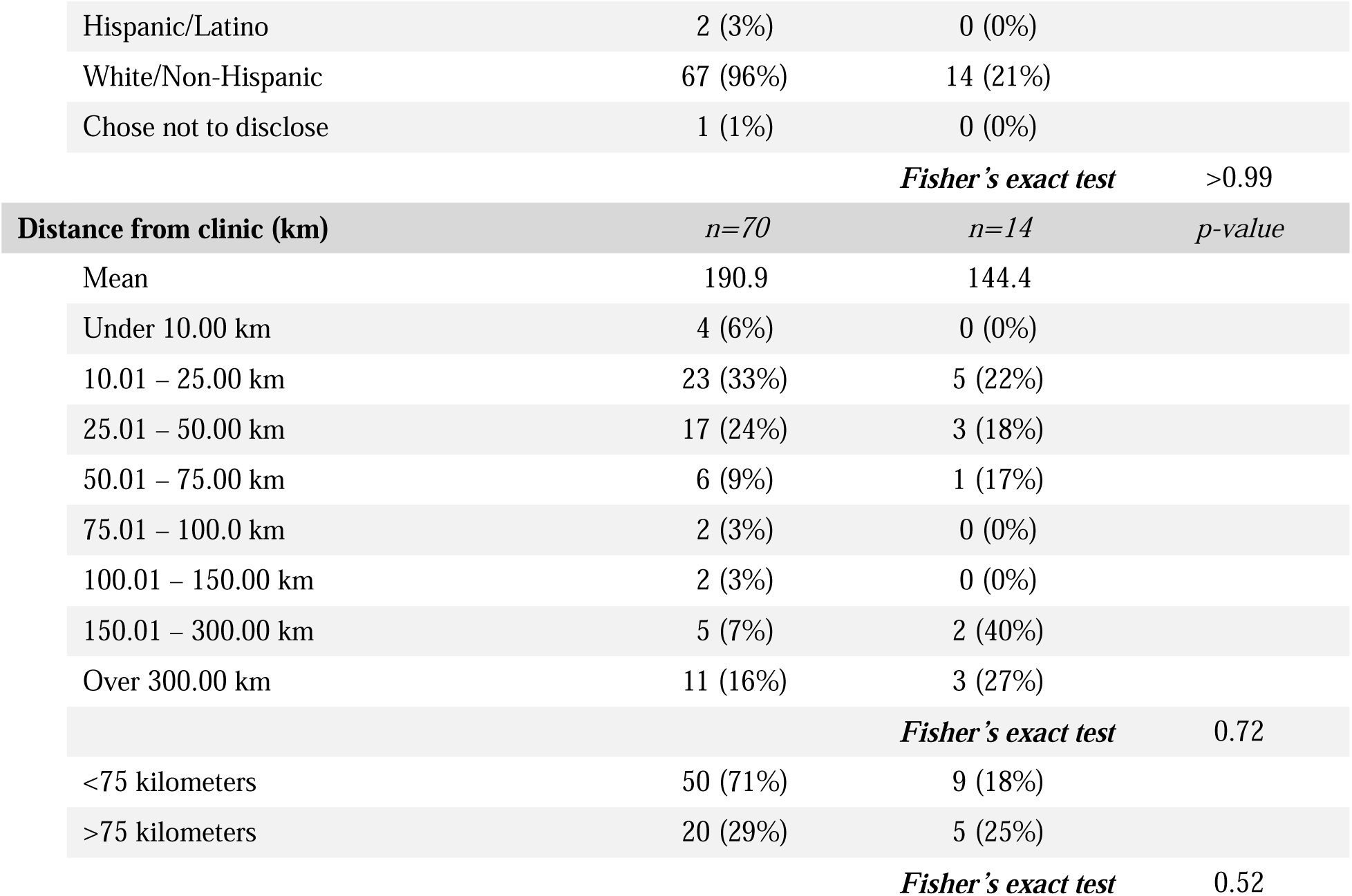
Patient demographics. Total patients and patients with ARIA incidence were stratified into various demographic groups, including age at different time points, sex, race/ethnicity, and distance from the clinical center. Percentages in the total column represent the portion of the total (n=70), while percentages within the ARIA cases column reflect the percentage of cases within each stratified sub-group/row. P-values were calculated using either Fisher’s exact test or Welch’s t-test, as noted. Values following the “±” symbol represent standard error of the mean (SEM).

On average, patients lived 190.9 kilometers the clinical center, with 20 out of 70 patients living greater than 75 kilometers from the clinical center. The furthest patient lived 3375.5 kilometers from our clinic, though this patient received infusions locally while visiting children who live in the area. We examined ARIA rates by distance to the University of Utah and found no statistically significant association between distance from the clinic and ARIA incidence (of the 14 patients with ARIA, 64% lived within 75 kilometers and 36% greater than 75 kilometers away) (**Table 2**).

The patient population included those with zero (30%), one (51%), or two copies (19%) of ApoE ε4. As expected, we observed a significant association between ApoE genotype and ARIA rates, with 46% of 13 ApoE ε4 homozygous patients, 11% of 36 ApoE ε4 heterozygous patients, and 19% of 21 patients with no copies of ApoE ε4 developing ARIA (Fisher’s exact test, p=0.03) (**Table 3**).

**Table 3:**
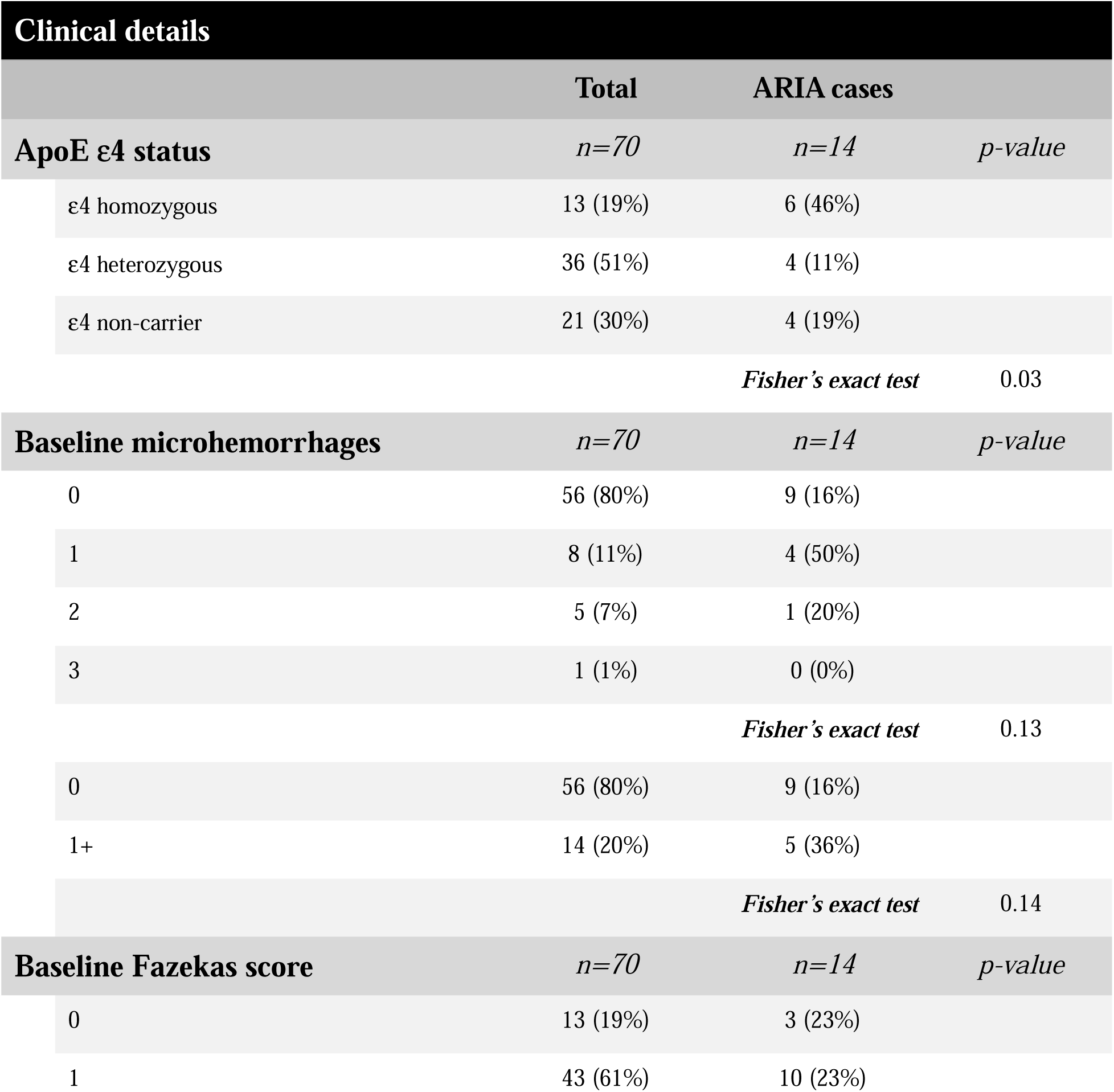
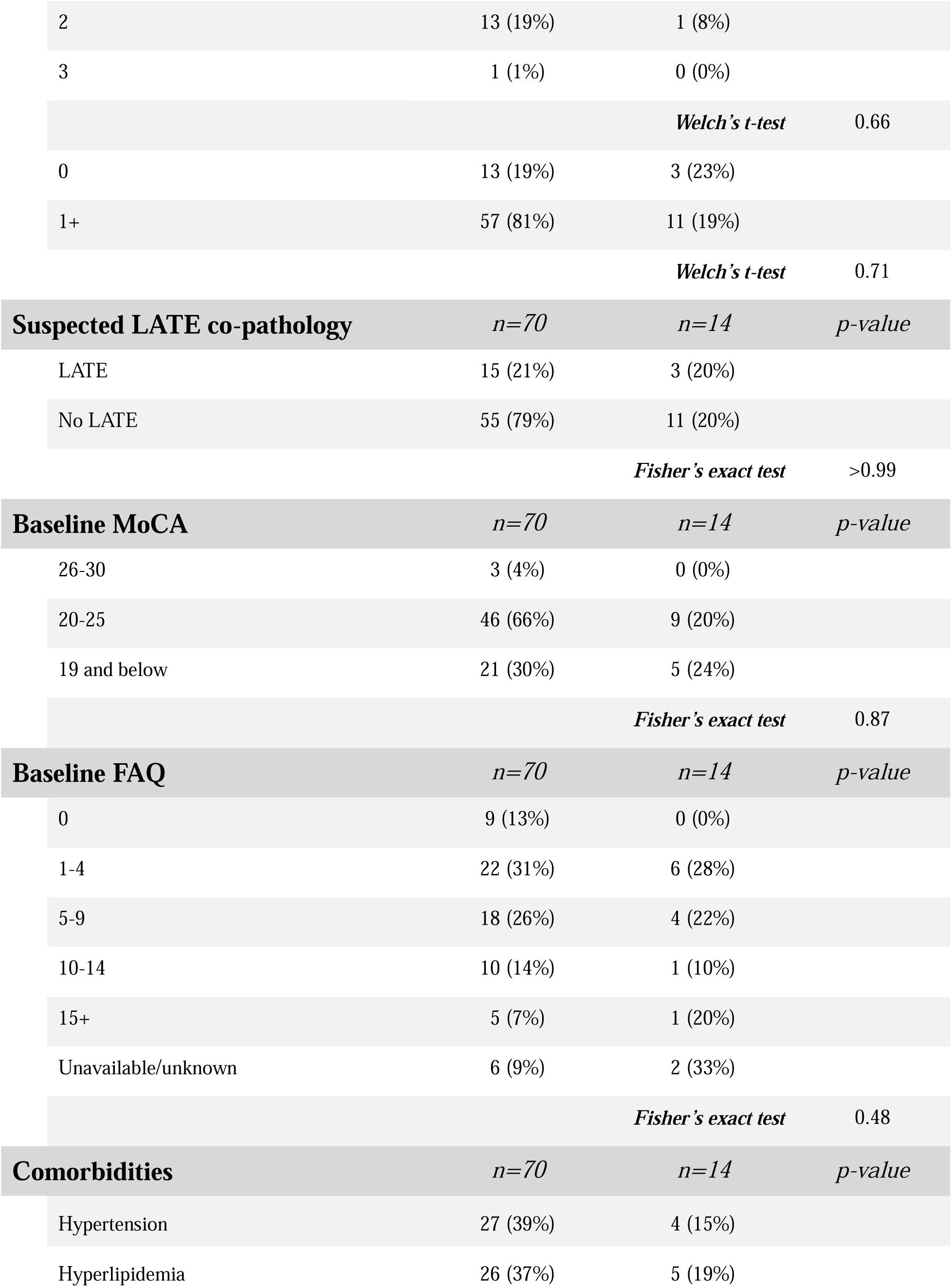
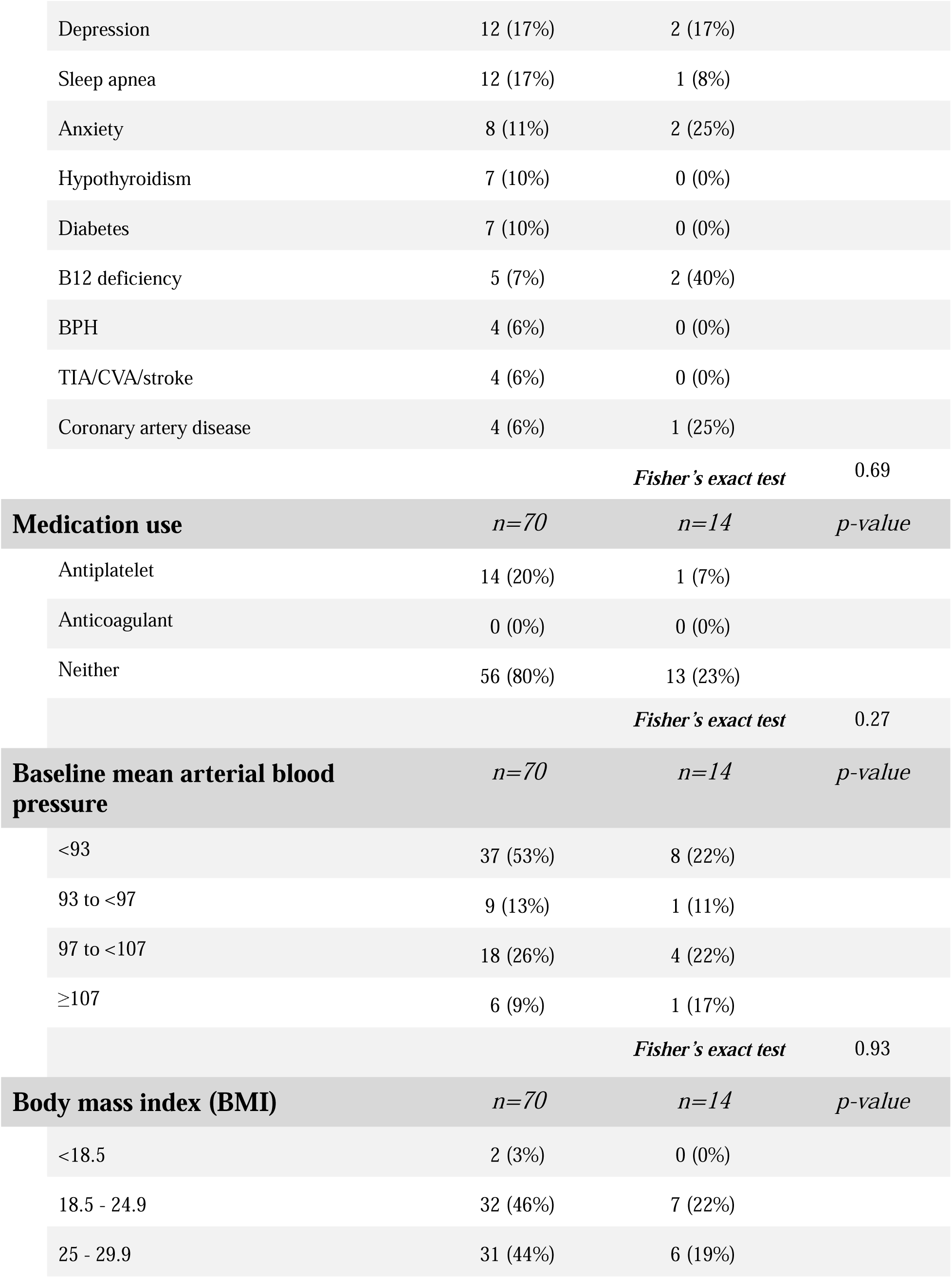
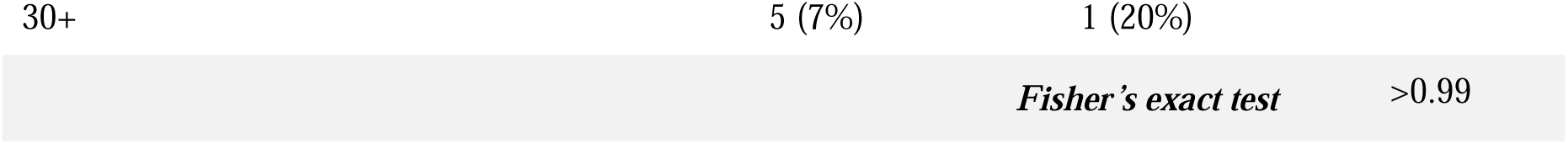
Clinical details. Total patients and patients with ARIA incidence were stratified according to ApoE ε4 status, as well as the number of microhemorrhages, Fazekas score found on baseline MRIs, patients with suspected LATE co-pathology, and baseline MoCA and FAQ scores. We also reviewed patient charts for comorbid health conditions, medication use, baseline mean arterial blood pressure, and BMI. Percentages of total represent the portion of the total (n=70), while percentages within the ARIA cases column reflect the percentage of cases within each stratified sub-group/row. P-values for each category were calculated using Fisher’s exact test and compare variability of stratified factors within the total and the ARIA cases group.

| **Clinical details** |  |  |  |
| --- | --- | --- | --- |
|  | **Total** | **ARIA cases** |  |
| **ApoE $\epsilon 4$ status** | *n=70* | *n=14* | *p-value* |
| $\epsilon 4$ homozygous | 13 (19%) | 6 (46%) | |
| $\epsilon 4$ heterozygous | 36 (51%) | 4 (11%) | |
| $\epsilon 4$ non-carrier | 21 (30%) | 4 (19%) | |
|  |  | *Fisher's exact test* | 0.03 |
| **Baseline microhemorrhages** | *n=70* | *n=14* | *p-value* |
| 0 | 56 (80%) | 9 (16%) |  |
| 1 | 8 (11%) | 4 (50%) |  |
| 2 | 5 (7%) | 1 (20%) |  |
| 3 | 1 (1%) | 0 (0%) |  |
|  |  | *Fisher's exact test* | 0.13 |
| 0 | 56 (80%) | 9 (16%) |  |
| 1+ | 14 (20%) | 5 (36%) |  |
|  |  | *Fisher's exact test* | 0.14 |
| **Baseline Fazekas score** | *n=70* | *n=14* | *p-value* |
| 0 | 13 (19%) | 3 (23%) |  |
| 1 | 43 (61%) | 10 (23%) |  |
| 2 | 13 (19%) | 1 (8%) |  |
| 3 | 1 (1%) | 0 (0%) |  |
|  |  | **Welch's t-test** | 0.66 |
| 0 | 13 (19%) | 3 (23%) |  |
| 1+ | 57 (81%) | 11 (19%) |  |
|  |  | **Welch's t-test** | 0.71 |
| **Suspected LATE co-pathology** | **n=70** | **n=14** | **p-value** |
| LATE | 15 (21%) | 3 (20%) |  |
| No LATE | 55 (79%) | 11 (20%) |  |
|  |  | **Fisher's exact test** | >0.99 |
| **Baseline MoCA** | **n=70** | **n=14** | **p-value** |
| 26-30 | 3 (4%) | 0 (0%) |  |
| 20-25 | 46 (66%) | 9 (20%) |  |
| 19 and below | 21 (30%) | 5 (24%) |  |
|  |  | **Fisher's exact test** | 0.87 |
| **Baseline FAQ** | **n=70** | **n=14** | **p-value** |
| 0 | 9 (13%) | 0 (0%) |  |
| 1-4 | 22 (31%) | 6 (28%) |  |
| 5-9 | 18 (26%) | 4 (22%) |  |
| 10-14 | 10 (14%) | 1 (10%) |  |
| 15+ | 5 (7%) | 1 (20%) |  |
| Unavailable/unknown | 6 (9%) | 2 (33%) |  |
|  |  | **Fisher's exact test** | 0.48 |
| **Comorbidities** | **n=70** | **n=14** | **p-value** |
| Hypertension | 27 (39%) | 4 (15%) |  |
| Hyperlipidemia | 26 (37%) | 5 (19%) |  |

|  |  |  |
| --- | --- | --- |
| Depression | 12 (17%) | 2 (17%) |
| Sleep apnea | 12 (17%) | 1 (8%) |
| Anxiety | 8 (11%) | 2 (25%) |
| Hypothyroidism | 7 (10%) | 0 (0%) |
| Diabetes | 7 (10%) | 0 (0%) |
| B12 deficiency | 5 (7%) | 2 (40%) |
| BPH | 4 (6%) | 0 (0%) |
| TIA/CVA/stroke | 4 (6%) | 0 (0%) |
| Coronary artery disease | 4 (6%) | 1 (25%) |

*Fisher's exact test* 0.69
| Medication use | *n=70* | *n=14* | *p-value* |
| --- | --- | --- | --- |
| Antiplatelet | 14 (20%) | 1 (7%) |  |
| Anticoagulant | 0 (0%) | 0 (0%) |  |
| Neither | 56 (80%) | 13 (23%) |  |

*Fisher's exact test* 0.27
| Baseline mean arterial blood pressure | *n*=70 | *n*=14 | *p*-value |
| --- | --- | --- | --- |
| <93 | 37 (53%) | 8 (22%) |  |
| 93 to <97 | 9 (13%) | 1 (11%) |  |
| 97 to <107 | 18 (26%) | 4 (22%) |  |
| ≥107 | 6 (9%) | 1 (17%) |  |

*Fisher's exact test* 0.93
| Body mass index (BMI) | *n*=70 | *n*=14 | *p*-value |
| --- | --- | --- | --- |
| <18.5 | 2 (3%) | 0 (0%) |  |
| 18.5 - 24.9 | 32 (46%) | 7 (22%) |  |
| 25 - 29.9 | 31 (44%) | 6 (19%) |  |
| 30+ | 5 (7%) | 1 (20%) |  |
| *Fisher's exact test* |  |  | >0.99 |

Patients were eligible for lecanemab if they had four or fewer microhemorrhages on baseline imaging. Thirteen patients (19%) had one or more microhemorrhage, while 81% of patients had a Fazekas score of 1 or greater. We observed that the presence of any preexisting microhemorrhages showed a trend toward increased incidence of ARIA (16% of 56 patients with no microhemorrhages vs. 36% of 14 patients with one or more microhemorrhage; Fisher’s exact test, p=0.14) (**Table 3**).

Other factors were not predictive of ARIA in our small cohort. For instance, patients with small vessel disease (Fazekas score >0; 18% of 57 patients) did not have an increased risk of ARIA compared to patients with no white matter disease (Fazekas score = 0; 23% of 13 patients). Co-pathology was suspected in a number of patients based on molecular imaging or history. Sixteen patients (23%) demonstrated hypometabolism in the medial temporal lobes, inferior frontal lobe, and anterior temporal lobe, suggestive of LATE^31^, but this group did not show a higher risk of ARIA (**Table 3**).

The average baseline MoCA score was 21 (range 16 - 29), and the average baseline FAQ was 6 (range 0 - 19). The majority of patients (89.6%) had a BMI between 18.5 and 29.9. The average baseline MAP was 92.9 (SEM 1.2). Examination of medical records revealed substantial co-morbidities, with hypertension, hyperlipidemia, and depression being observed in 27 (39%), 26 (37%), and 12 (17%) patients, respectively (**Table 3**).

Neither baseline MoCA nor baseline FAQ showed significant associations with ARIA, though we did not observe any ARIA in the three patients with a MoCA >25 or the nine patients with an FAQ of 0. Similarly, hypertension (or baseline MAP), hyperlipidemia, depression, and other co-morbidities did not have a clear relationship with ARIA rates, and antiplatelet use also did not raise ARIA risk, consistent with prior studies ^19,34^ (**Table 3**).

We next examined whether CSF analytes would predict ARIA rates, as a total of 53 of our patients underwent CSF analysis for confirmation of amyloid pathology. Of these patients, five underwent testing using version 1 of the quantiferon test. We examined the reported CSF p-tau181/Aβ42 ratio, Aβ42, t-tau, and p-tau in the 48 patients who were tested with version 2. Of these, eight patients developed ARIA and 40 patients did not. Although there were on average lower Aβ, higher tau, and elevated ratios in patients who developed ARIA, none of these factors reached significance (**Table 4**).

**Table 4:** Biomarkers—CSF Aβ and tau are not predictive of ARIA risk. Out of 70 total patients, 53 received lumbar puncture and cerebral spinal fluid testing to confirm AD diagnosis. Of those 53, five received an earlier version of the testing not detailed here. The average ratio of p-tau181/Aβ42, total Aβ42, total tau, and total p-tau of the remaining 48 patients with the more recent test are included here in total, as well as divided by ARIA incidence and no ARIA incidence. The included p-values are calculated for each diagnostic average between the ARIA case and no ARIA groups using a two-way ANOVA with Bonferroni’s multiple comparisons test. Values following the “±” symbol represent standard error of the mean (SEM).

| Biomarkers |  |  |  |  |
| --- | --- | --- | --- | --- |
|  | Total | ARIA cases | No ARIA |  |
| **Average CSF ratio** | *n*=48 | *n*=8 | *n*=40 | *p*-value |
| Reference range $\leq 0.028$ | 0.058 ± 0.003 | 0.067 ± 0.006 | 0.056 ± 0.004 | |
| *Two-way ANOVA with Bonferroni's multiple comparisons* |  |  |  | 0.48 |
| **Average Aβ42** | *n*=48 | *n*=8 | *n*=40 | *p*-value |
| Reference range $> 834$ | 702 ± 41 | 656 ± 89 | 712 ± 43 | |
| *Two-way ANOVA with Bonferroni's multiple comparisons* | | | | $> 0.99$ |
| **Average CSF tau** | *n*=48 | *n*=8 | *n*=40 | *p*-value |
| Reference range $\leq 238$ | 358 ± 21 | 403 ± 52 | 349 ± 22 | |
| *Two-way ANOVA with Bonferroni's multiple comparisons* |  |  |  | 0.80 |
| **Average p-tau** | *n*=48 | *n*=8 | *n*=40 | *p*-value |
| Reference range $\leq 21.6$ | 38.8 ± 2.5 | 42.8 ± 5.6 | 37.9 ± 2.8 | |
| *Two-way ANOVA with Bonferroni's multiple comparisons* | | | | $> 0.99$ |

Initiation of therapy following the decision to start is a complex process involving prior authorization with insurance and scheduling with infusing centers. Similarly, completion of infusion courses involves logistical hurdles of scheduling infusions, surveillance MRIs, and follow-up appointments, as well as patient-related life events (illness, travel, etc.).

As of July 1, 2025, the majority of patients (61%) had completed 13 or more infusions. Patients who were local received infusions from university infusion centers, whereas patients in more distant locations were assisted in arranging infusions in their respective areas. A total of 15 patients received infusions at non-university infusion sites, all of whom lived more than 75 kilometers from the University of Utah, with an average of 352.1 kilometers from the clinical center. We found that patients located within 75 kilometers and those greater than 75 kilometers had similar onset to initiation of therapy and to completion of 13 or 26 doses of lecanemab (**Table 5**). A total of 13 patients (19%) discontinued therapy for any reason. There was no relationship between the distance to the University of Utah and discontinuation of therapy. Twelve patients (92%) of the 13 who discontinued therapy were within 75 kilometers of the University of Utah (**Table 5**).

**Table 5:**
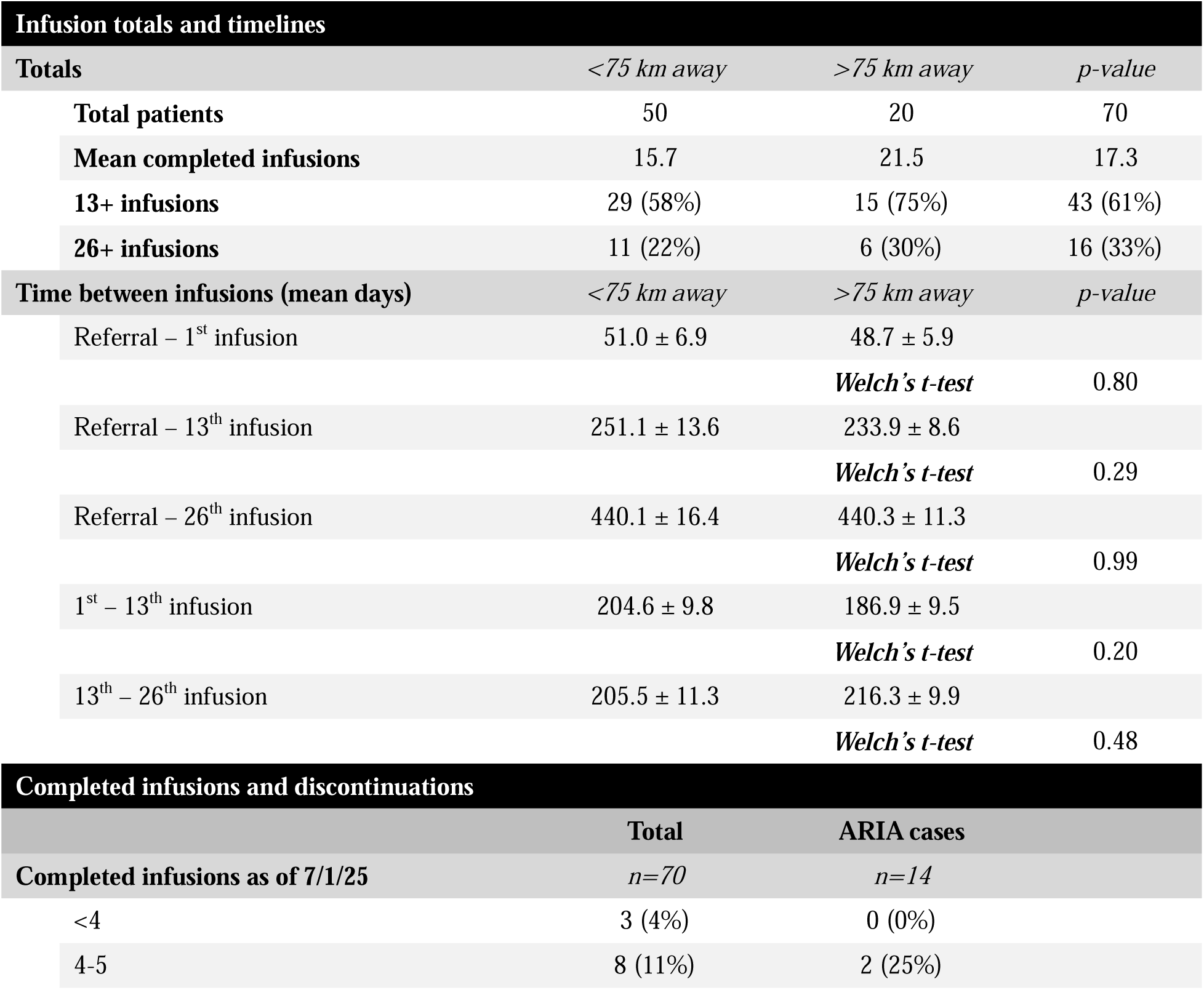
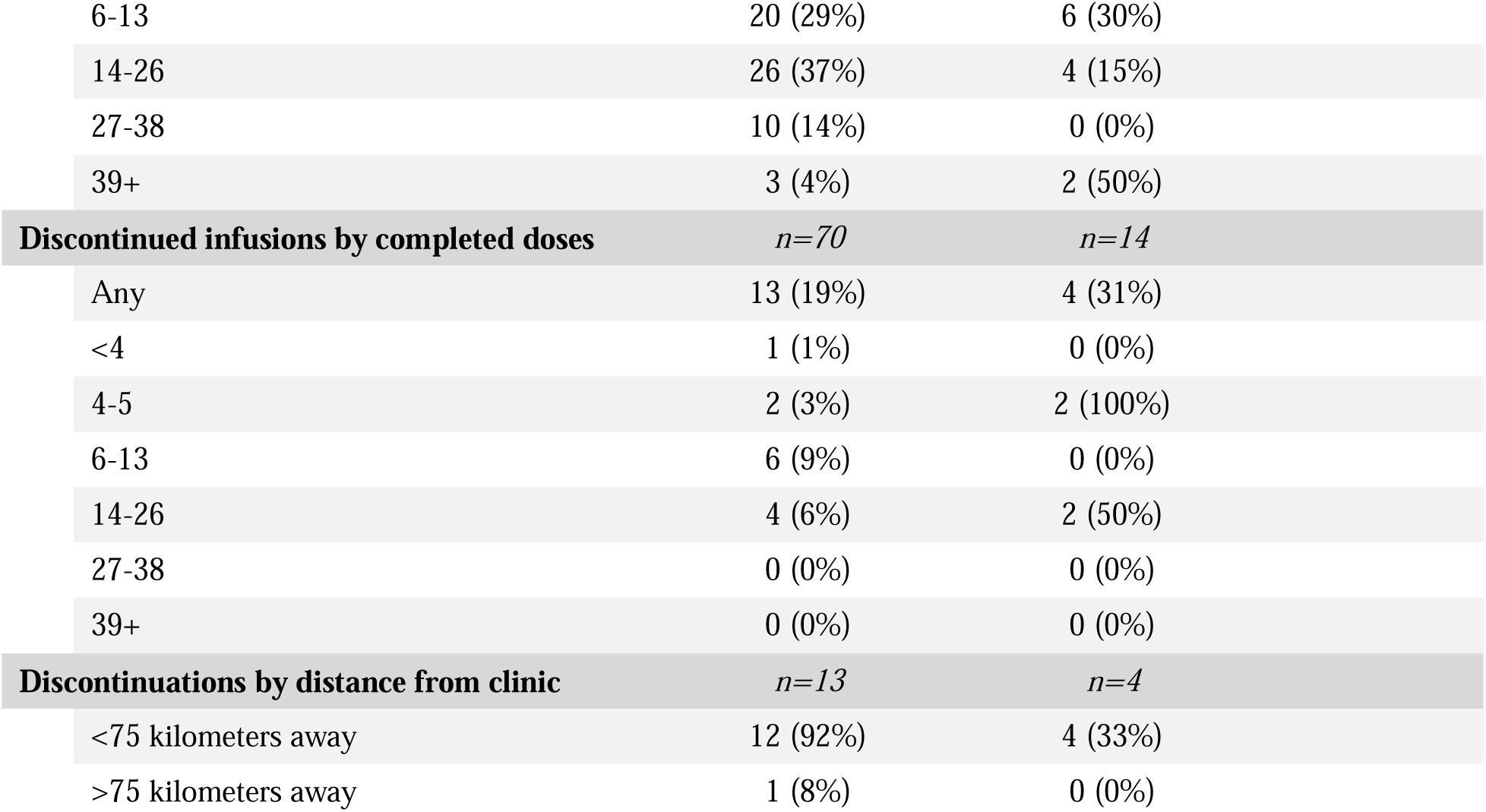
Infusion details. Patients were segmented according to distance of home address from the clinical center. This table details infusion totals and timelines between infusions based on distance from the clinic P-values were calculated between the two groups using Welch’s *t*-test. Values following the “±” symbol represent standard error of the mean (SEM).. Total patients and patients with ARIA are also detailed according to completed infusions as of 7/1/25, as well as patients who discontinued infusions for any reason. For completed infusions and discontinued infusions, percentages of the total represent the portion of the total (n=70), while percentages within the ARIA cases column reflect the percentage of cases within each sub-group/row. Among discontinuations by distance from clinic, percentages of the total include only those patients who discontinued therapy (n=13), while the ARIA cases column reflects the percent of discontinuations within each geographic group that experienced ARIA.

| Infusion totals and timelines |  |  |  |
| --- | --- | --- | --- |
| Totals | <75 km away | >75 km away | p-value |
| Total patients | 50 | 20 | 70 |
| Mean completed infusions | 15.7 | 21.5 | 17.3 |
| 13+ infusions | 29 (58%) | 15 (75%) | 43 (61%) |
| 26+ infusions | 11 (22%) | 6 (30%) | 16 (33%) |
| Time between infusions (mean days) | <75 km away | >75 km away | p-value |
| Referral – 1<sup>st</sup> infusion | 51.0 ± 6.9 | 48.7 ± 5.9 |  |
|  |  | *Welch's t-test* | 0.80 |
| Referral – 13<sup>th</sup> infusion | 251.1 ± 13.6 | 233.9 ± 8.6 |  |
|  |  | *Welch's t-test* | 0.29 |
| Referral – 26<sup>th</sup> infusion | 440.1 ± 16.4 | 440.3 ± 11.3 |  |
|  |  | *Welch's t-test* | 0.99 |
| 1<sup>st</sup> – 13<sup>th</sup> infusion | 204.6 ± 9.8 | 186.9 ± 9.5 |  |
|  |  | *Welch's t-test* | 0.20 |
| 13<sup>th</sup> – 26<sup>th</sup> infusion | 205.5 ± 11.3 | 216.3 ± 9.9 |  |
|  |  | *Welch's t-test* | 0.48 |
| Completed infusions and discontinuations |  |  |  |
|  | Total | ARIA cases |  |
| Completed infusions as of 7/1/25 | n=70 | n=14 |  |
| <4 | 3 (4%) | 0 (0%) |  |
| 4-5 | 8 (11%) | 2 (25%) |  |

|  |  |  |
| --- | --- | --- |
| 6-13 | 20 (29%) | 6 (30%) |
| 14-26 | 26 (37%) | 4 (15%) |
| 27-38 | 10 (14%) | 0 (0%) |
| 39+ | 3 (4%) | 2 (50%) |
| **Discontinued infusions by completed doses** | *n=70* | *n=14* |
| Any | 13 (19%) | 4 (31%) |
| <4 | 1 (1%) | 0 (0%) |
| 4-5 | 2 (3%) | 2 (100%) |
| 6-13 | 6 (9%) | 0 (0%) |
| 14-26 | 4 (6%) | 2 (50%) |
| 27-38 | 0 (0%) | 0 (0%) |
| 39+ | 0 (0%) | 0 (0%) |
| **Discontinuations by distance from clinic** | *n=13* | *n=4* |
| <75 kilometers away | 12 (92%) | 4 (33%) |
| >75 kilometers away | 1 (8%) | 0 (0%) |

Initial visits were in-person encounters; however, on subsequent visits, patients could elect for telehealth visits. Telehealth options were utilized by patients regardless of their distance from the clinical center, with a slightly larger proportion of more remote patients (located >75 kilometers away) utilizing one or more telehealth visit—78% vs. 70% of locals. While there was no difference between these two groups in the number of patients who were seen in follow-up visits or completed at least one virtual visit, there was a trend toward greater use of virtual visits among patients who were >75 kilometers from the University of Utah, with a larger proportion of telehealth visits out of total follow-up visits in this group—58% vs. 42% among locals (Table 6). DISCUSSION

**Table 6:** Follow-up visits. Regular follow-up visits are a part of lecanemab treatment, and many patients opt for virtual visits with the provider as opposed to in-person appointments. Here we break down patients by distance from the clinic and include follow-up visit totals and relative proportion of virtual visits. Percentages of total with follow-up visits are out of the total patients within each group (<75 kilometers vs. >75 kilometers away). Percentages of those with 1+ virtual follow-up are a portion of those with any follow-up visits since starting treatment. P-values were calculated using Fisher’s exact test.

| Follow-up visits |  |  |  |
| --- | --- | --- | --- |
|  | <75 km away | >75 km away | p-value |
| Total patients | 50 | 20 |  |
| Total with follow-up visits | 43 (86%) | 18 (90%) |  |
|  |  | *Fisher's exact test* | >0.99 |
| 1+ virtual follow-up visit | 30 (70%) | 14 (78%) |  |
|  |  | *Fisher's exact test* | 0.59 |
| Total visits | 124 | 59 |  |
| Total virtual visits | 52 (42%) | 34 (58%) |  |
|  |  | *Fisher's exact test* | 0.057 |

Here we present real-world data on patients with mild cognitive impairment or mild dementia due to Alzheimer’s disease who were treated with lecanemab at the University of Utah. In total we report on 70 patients who were evaluated in our clinical center and who received at least one dose of lecanemab. Similar to prior real-world studies^32-34^, we find that lecanemab is generally safe and well tolerated and that side effects are in line with those reported in large, controlled trials^7,17,20,35,36^. Moreover, while patient evaluation, drug administration, and monitoring for adverse events each present significant logistical challenges, we demonstrate that lecanemab administration can be safely performed in distributed areas, aided by telehealth tools.

Overall, we observed a rate of ARIA (20%) that is similar to those found in prior real-world data and controlled trials^7,20,24,32-34^. As expected, ApoE genotype is significantly associated with development of ARIA. Moreover, controlled studies have repeatedly demonstrated that ARIA occurs in a ApoE-dependent fashion even in the absence of amyloid-removing therapies^7,17,22,37^. This highlights that ARIA is strongly associated with amyloid deposition regardless of treatment. While other studies report that the existence of prior microhemorrhages also increases ARIA risk^7,17,33^, our analysis showed only a trend in this direction. Among patients with existing microhemorrhages on baseline susceptibility imaging, the proportion of ARIA incidence was nearly twice as high than among those with no baseline microhemorrhages. We had no symptomatic cases of ARIA.

We also examined the effect of other comorbidities in driving risk for ARIA. Notably, although we had high levels of comorbid cardiovascular risk factors, these were lower than those reported in other studies, which may be related to our method of retrospective chart analysis. A larger percentage of our patients received metabolic PET imaging, allowing us to identify patients with probable TDP-43 co-pathology (LATE) based on hypometabolism in medial temporal lobes and frontal lobes^31,38-40^. Patients with suspected co-pathology were counseled that amyloid-removal therapy would likely not alter TDP-43 pathology; however, it was not expected that TDP-43 pathology would increase risk of treatment. Our data supports this, as we observe similar rates of ARIA in patients with and without suspected LATE, though notably we do not have biopsy confirmation of TDP-43 pathology.

Our analysis supports the effectiveness and safety of treating patients across a widely distributed area. While the majority of patients (71%) lived within 75 kilometers of the clinical center, the 29% located farther out showed no significant difference in treatment progression or outcomes—both in terms of ARIA incidence as well as infusion timelines (i.e., time from referral to starting and continuing infusions). This was likely facilitated by greater utilization of virtual visits in this group.

There are some limitations to our study. Most notably, the sample size was relatively small, which may limit the statistical power and generalizability of our findings. Additionally, as the clinic is located in a region with limited demographic diversity, the study population reflects the broader, largely White patient population. Future research in other locales that could include larger, more diverse cohorts would be useful to validate these findings and assess their relevance across more wide-ranging demographic and geographic contexts.

In this report we have focused on our initial experience with lecanemab. With the more recent FDA approval of once-monthly donanemab^17^ and more recent data suggesting that modified titration of donanemab improves safety^41^, it is expected that amyloid-removal therapies will become more feasible for a greater number of patients located outside of the vicinity of large academic medical centers. We expect our data will help to alleviate concerns about safety as patients are treated in increasingly less well-controlled settings.

## Data Availability

Data presented in this manuscript may be shared with approval of all appropriate Institutional Review Boards.

## ACKNOWLEDGEMENTS

The authors confirm their contribution to this work as follows. N.A.F. and B.C.C. wrote the paper with input from all authors. N.A.F. and B.C.C., contributed to the conception and design of the study. Patients were adjudicated by N.A.F., C.J.C., M.K.S., Y.A., and S.M. Data was collected by B.C.C. and C.Z., and analyzed by B.C.C. under the supervision of N.A.F. Funding for the study was obtained by N.A.F., A.P. and C.J.C.

## CONSENT STATEMENT

Informed consent was waived in this retrospective study. All work was overseen and approved by the University of Utah Institutional Review Board. This study was conducted in accordance with the principles of the Declaration of Helsinki.

---

**Supplementary table 1:**
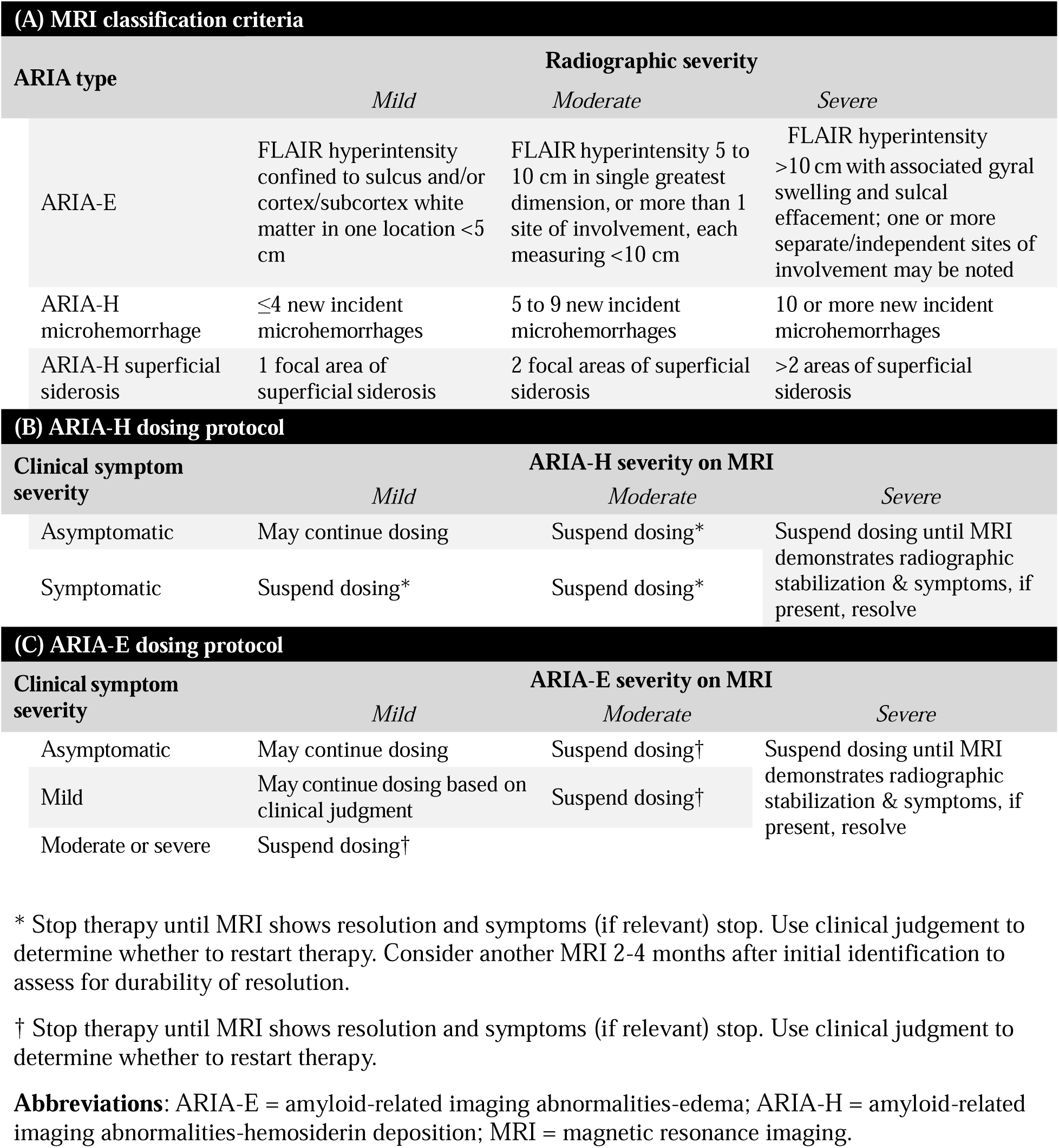
ARIA MRI classification and dosing protocol. Incident ARIA-E and ARIA-H in our patient population was evaluated using the following criteria (Adapted from Cummings et al, 2023) ^24^:

